# Association Between Purchase of Organic Food and Hypertension Among US Adults: NHANES 2007-2010

**DOI:** 10.64898/2026.05.13.26353146

**Authors:** Christopher Choi, Liwei Chen, Onyebuchi A. Arah, Roch A. Nianogo

## Abstract

**Background:** An increasing demand for organic food has risen due to perceived health benefits. Current evidence for the health effects of organic food is limited.

**Objective:** To evaluate the association between organic food purchase as a proxy for organic food consumption and hypertension in a nationally representative population of the US.

**Methods:** This was a cross-sectional study that included 9173 participants aged ≥ 18 and had available data of both organic food purchase and hypertension from the National Health and Nutrition Examination Survey 2007-2010. Organic food purchase and frequency were obtained from survey questionnaires. Hypertension was defined as having either a systolic BP ≥ 130 mm Hg/ diastolic BP ≥ 80 mm Hg, currently taking antihypertensive medication, or self-reported diagnosis of hypertension. We used multivariable logistic regression with sample weights and adjustment of potential confounders to assess associations (adjusted odds ratio [aOR] and 95% confidence intervals [CI]) between organic food purchase and hypertension status.

**Results:** Findings suggest an 11% decrease in odds of hypertension (aOR = 0.89, 95% CI: 0.75– 1.06) among organic food purchasers compared to non-purchasers. Lower odds of hypertension were observed across all categories of organic food purchasing frequency, with 13% lower among rarely purchasing organic food (aOR = 0.87, 95% CI: 0.67–1.14), 9% lower (aOR = 0.91, 95% CI: 0.71–1.16) among sometimes purchasing organic food, and 17% lower (aOR = 0.83, 95% CI: 0.55–1.27) among always or mostly purchasing organic food, as compared to those who never purchased organic food.

**Conclusion:** Our findings suggest that organic food purchase, a proxy for organic food consumption, may be associated with lower odds of hypertension. These findings may reflect either the true benefits of organic food consumption, including lower pesticide amounts and higher nutrient content, or the health-seeking behaviors among health-conscious, healthy, and highly educated individuals.

## 1. Introduction

Hypertension is a major modifiable risk factor for cardiovascular disease (CVD) and premature death worldwide ^1^. In 2024, it was estimated that 1.4 billion adults aged 30-79 years old had hypertension globally ^2^. In the United States alone, 119.9 million adults had hypertension, which accounts for nearly half the US adult population ^3^. Hypertension has been extensively studied in the past century, and accumulating evidence highlights the importance of modifiable risk factors in reducing its occurrence. These modifiable risk factors include an unhealthy diet, inactive lifestyle, smoking, and consumption of alcohol ^2,4^. Making lifestyle changes not only reduces the risk of hypertension, but also results in a 15% reduction of cardiovascular-related events ^4^.

Diet is one of the key modifiable risk factors for hypertension, and one widely recognized diet approach is the Dietary Approaches to Stop Hypertension (DASH). The DASH diet plan recommends eating fruits, vegetables, and whole grains; low-fat and/or fat-free dairy products, poultry, fish, beans, nuts, and vegetable oil; and limiting food in saturated fat, trans fat, sweets, and sugar-sweetened beverages ^5^. The DASH diet has been shown to reduce blood pressure (BP) levels, even in individuals without hypertension ^6–9^. While the DASH diet focuses on selecting specific food groups, the agricultural methods used to produce those foods may further influence BP through organic farming practices.

Organic foods incorporate strict farming practices that do not allow genetically modified organisms, no synthetic pesticides or chemical fertilizers, no antibiotics or growth hormones, and avoid processing through industrial solvents or chemical food additives^9,10^. The absence of these practices in organic foods may confer health benefits that conventional foods may not. Organic foods are nutrient-dense with higher levels of omega-3 fatty acids in dairy products, increased levels of antioxidants in organic crops, and improved fatty acid levels in meat products ^11–14^. Also, compared to conventionally grown crops, organically grown crops have one-third the amount of pesticide residue detected ^12,15^. To date, few studies have examined the indirect health effects of organic food on hypertension. The KOALA birth cohort study found that organic food consumers had lower hypertension incidence during pregnancy, though the association with blood pressure was not linear ^16^. Similarly, the NutriNet-Santé study reported that higher consumption of organic foods was associated with lower odds of metabolic syndrome, which included elevated blood pressure as a diagnostic criterion ^17^. However, studies have investigated the relationship between organic food consumption and other cardiometabolic health outcomes, including obesity, metabolic syndrome, atherosclerotic cardiovascular disease, and diabetes. Results from these studies suggest organic food consumption may be associated with a lower likelihood of adverse health outcomes ^17–20^.

Potential mechanisms underlying the effects of organic food on blood pressure include the possibility of reducing pesticide exposure and increasing health-beneficial nutrient intake. In a randomized controlled trial, a diet rich in polyphenol-rich juices reduces blood pressure in both normal and hypertensive individuals ^21^. Another study found that a higher intake of antioxidants resulted in lower systolic blood pressure ^22^. A dose-response relationship of omega-3 fatty acids demonstrated that an optimal intake between 2 g/d and 3 g/d lowered blood pressure ^23^. In the US, the most commonly used insecticides are organophosphates, pyrethroids, and carbamates. Organophosphates, among them, are widely used in agriculture, and humans consume them ^24^. One plausible mechanism by which organophosphates may contribute to hypertension is by inhibiting acetylcholinesterase (AChE). Inhibition of AChE disrupts cholinergic pathways that regulate blood pressure and cardiovascular function ^25,26^. Studies of the NHANES 2013-2014 and 2015-2016 cycles found that pesticide exposure is associated with blood pressure dysregulation, as measured by urinary and blood biomarkers. However, these associations appear to be metabolite dependent, meaning they vary according to the specific breakdown products of organophosphate insecticides ^25,27^. Overall, the evidence is mixed, suggesting that some metabolites raise BP, while others lower it.

Although individual mechanisms of organic food have been explored in relation to blood pressure, it remains unclear whether these mechanisms, when considered together, contribute to a measurable association between organic food and hypertension. The objective of this study was to investigate the association between the purchase of organic food and the frequency of organic food purchases (as a proxy for organic food consumption) and hypertension.

## 2. Materials and Methods

### 2.1. Study Population

The study population is drawn from the 2007-2010 cycles of the National Health and Nutrition Examination Survey (NHANES). NHANES is a cross-sectional survey that collects data from a representative sample of the U.S. civilian noninstitutionalized population by using complex, multistage probability clusters. The data collected consists of health and nutrition information of adults and children within communities across the United States. Questionnaire data were collected through face-to-face interviews conducted by a trained interviewer at participants’ homes. Health examination data were collected using a mobile examination center (MEC) that travels across the country to obtain a range of measurements, such as blood, urine, and blood pressure. In the 2007-2010 NHANES cycles, participants were aged 18 and over. Participants excluded were those without information on organic food purchases. A total of 9173 participants were included in the analysis. Data on the frequency of organic food purchases were available for 7426 participants, as this question was only administered during the 2007-2008 cycle.

### 2.2. Exposures: Organic food purchase and frequency of organic food purchase

Consumer behavior was recorded, and participants were asked about their primary exposure: the purchase of organic foods. Participants were asked, “In the past 30 days, did you buy any food that had the word ‘organic’ on the package?” Purchase of organic food was defined as “Yes” and “No”.

Secondary exposure captured the frequency of organic food bought. Participants were asked, “How often do you buy organic food? Would you say always, most of the time, sometimes, or rarely?” Those who responded “Always” and “Most of the time” were combined and collapsed into a single response, labeled “Always/Most”. This was done because the sample size was smaller than that of the other responses.

### 2.3. Outcomes: Hypertension

Blood Pressure (BP) readings were measured at the mobile examination center by certified BP examiners who had completed and passed a training program from Shared Care Research and Education Consulting. The procedure to measure participants’ BP was to have them quietly sit for 5 minutes and determine the maximum inflation level. After completing these 2 requirements, three consecutive blood pressure readings were then measured. If a reading was incomplete or interrupted, a 4^th^ measurement was recorded. All blood pressure readings were recorded as systolic and diastolic measurements in mm Hg. Only the 2^nd^ and 3^rd^ BP readings were used to calculate a single average BP measurement. The 1^st^ reading was excluded due to the possibility of white coat hypertension. This is when a blood pressure reading is higher than normal when measured in a healthcare setting. The condition is likely caused by anxiety when in a health care setting, and the presence of a health care provider measuring their BP. Hypertension was defined as having either a systolic BP ≥ 130 mm Hg or a diastolic BP ≥ 80 mm Hg, or currently taking antihypertensive medication, or self-reported diagnosis of hypertension. Hypertension was categorized into Yes and No. In the sensitivity analysis, the BP threshold was raised to a systolic BP ≥ 140 mm Hg or a diastolic BP ≥ 90 mm Hg, consistent with the previously established hypertension guidelines.

### 2.4. Covariates and potential confounders

#### Sociodemographic variables

gender (female, male), age (continuous in years), race, education, and the ratio of family income to poverty were obtained during the interview. Race was categorized as Mexican American, Hispanic, Non-Hispanic White, Non-Hispanic Black, and Other Race – Including Multi-Racial. Mexican American and Hispanic were combined into a single Hispanic category, resulting in 4 categories: Hispanic, White, Black, and Other. Education levels ranging from less than 9th grade to college graduate or above, with 5 levels, were categorized into 3. They are less than high school, high school/GED equivalent, and college or higher. The family-income-to-poverty ratio was categorized as <1.00, 1.00-1.99, 2.0-3.99, and >4.00.

#### Lifestyle factor variables

Smoking, alcohol, physical activity, body mass index (BMI), and dietary intake (total energy intake & overall diet quality) were also obtained during the interview and in the MEC. Smoking status for individuals who smoked less than 100 cigarettes in their lifetime was defined as never smokers; those who smoked more than 100 cigarettes and smoked at the time of the survey were defined as current smokers; those who had smoked more than 100 cigarettes but did not smoke during the time of the survey were defined as former smokers. Alcohol use was categorized into non-drinker, moderate, and heavy. Alcohol use was defined as having drunk any alcohol within the past 12 months at the time of the survey. Individuals who had 0 drinks were defined as non-drinkers, men who had 2 drinks or less in a day and women who had 1 drink or less in a day were defined as moderate drinkers, and men who had 15 or more drinks in a week and women who had 8 or more drinks in a week were defined as heavy drinkers. Physical activity (PA) was assessed using the Global Physical Activity Questionnaire. PA was defined as the combination of total vigorous and moderate weekly minutes of domain-specific physical activity. The domains are occupational, recreational, and transportation PA. To have a standardized unit of minutes, vigorous minutes were multiplied by 2 to obtain moderate-equivalent minutes. Total weekly minutes grouped into 0 min, 1–149 min, 150–299 min, 300–599 min, and 600 + minutes. The weekly minutes were defined as at least 150 minutes of moderate intensity or 75 minutes of vigorous intensity activities per week. Body mass index (BMI) data were collected by health technicians at the MEC. BMI was calculated by dividing a person’s weight in kilograms by the square of their height in meters. Dietary intake was assessed as the average of two 24-hour dietary recalls. Total energy intake was defined as total kcal consumed. Overall diet quality was assessed using the Healthy Eating Index 2015 (HEI) and is scored based on the intake levels of 13 dietary components, including total fruit, whole fruit, total vegetables, greens and beans, whole grains, dairy, total protein foods, seafood and plant proteins, fatty acids, refined grains, sodium, added sugars, and saturated fats. Scores range from 0 to 100, with higher scores indicating a good diet.

### 2.5. Statistical analysis

The complex, multistage, stratified, cluster-sampling design of NHANES was accounted for in all analyses using sample weights, strata, and primary sampling units. Missing data were handled by excluding observations with missing exposure and outcome information at baseline. Remaining missing values were addressed using multiple imputation by chained equations with M = 10 imputed datasets. Before running the imputation, logical constraints were defined to prevent imputing missing values for the frequency of organic food purchase among 2009-2010 cycle participants, due to structural missingness. Multivariable logistic regression models were used to estimate the association between hypertension and organic food purchase and its frequency, adjusting for confounders. In a partially adjusted model (model 1), age, gender, race, education, and family-income-to-poverty ratio were included. For the fully adjusted model (model 2), smoking status, alcohol intake, total physical activity, BMI, total energy intake, and HEI score were included. After running the logistic regression models, the estimates from each imputed data set were pooled together to produce one single estimate for each model.

We conducted a sensitivity analysis to assess the odds of hypertension by raising the BP threshold to a systolic BP ≥ 140 mm Hg or diastolic BP ≥ 90 mm Hg, consistent with the previously established hypertension guidelines. Statistical analysis was conducted using R, version 4.5.2 (R Foundation for Statistical Computing, Vienna, Austria).

## 3. Results

### 3.1. Demographic characteristics

A total of 9173 participants were included in this study, with a mean age of 4 years (SD = 18). Participants who purchased food in the last 30 days were more likely to be female, White, college-educated, with a higher income-to-poverty ratio, non-smokers, moderate drinkers, and higher levels of physical activity. BMI categories were similar in the purchase of organic food (**Table 1**).

**Table 1.**
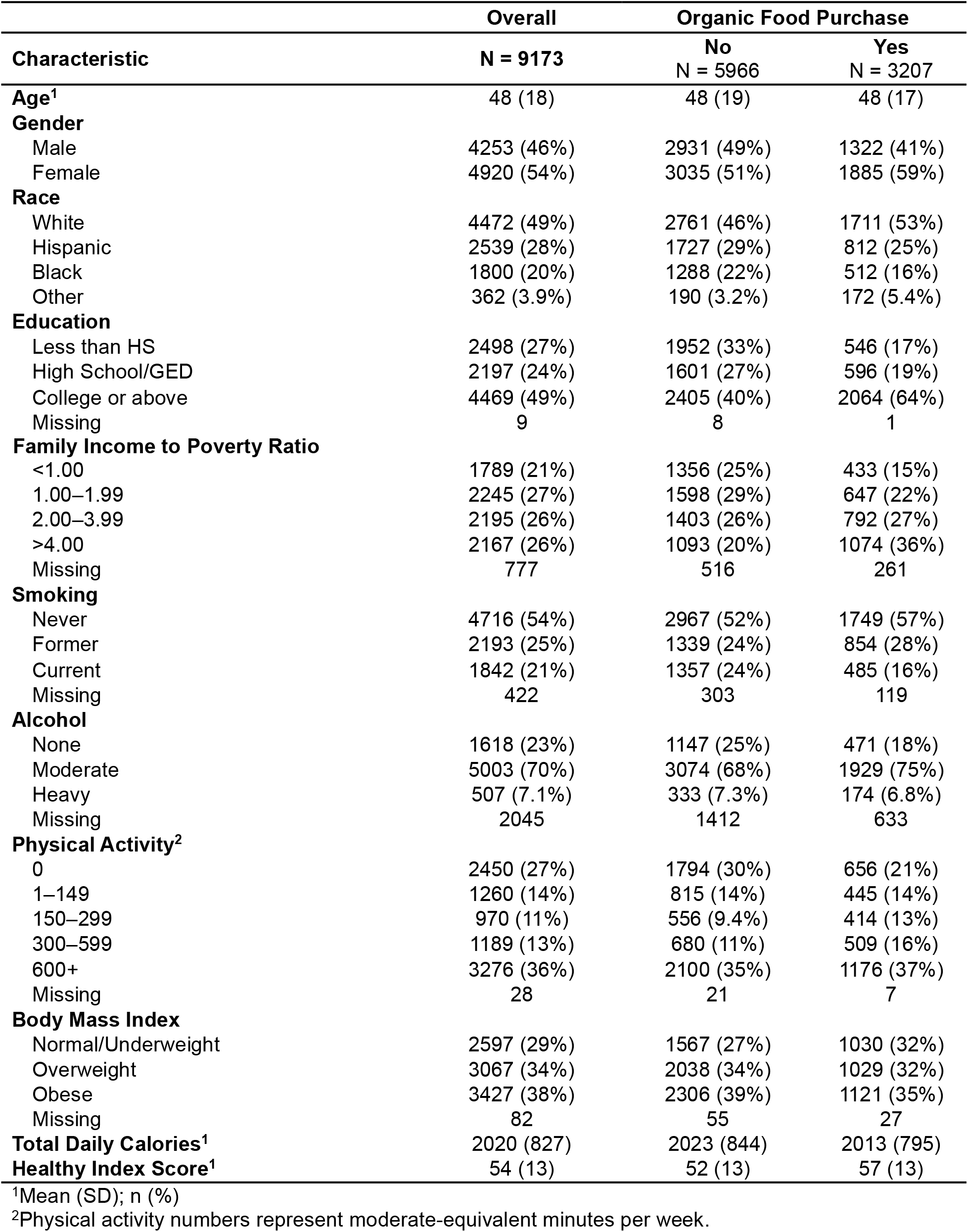
Baseline characteristics of participants in NHANES 2007–2008, according to their purchase of organic foods.

### 3.2. Association of organic food purchase with hypertension

The purchase of organic food was associated with a 11% decrease in the odds of hypertension. An odds ratio ranging from a 25% decrease, a moderate negative association, to a 6% increase, a minimal positive association, is reasonably compatible (aOR = 0.89, 95% CI: 0.75–1.06) (**Table 2)**.

**Table 2.** Association between organic food purchase and hypertension, NHANES 2007–2008.

|  | **Odds Ratio (OR)**<br>**95% CI**<br>**Crude** | **Odds Ratio (OR)**<br>**95% CI**<br>**Model 1<sup>1</sup>** | **Odds Ratio (OR)**<br>**95% CI**<br>**Model 2<sup>2</sup>** |
| --- | --- | --- | --- |
| **Organic food purchase**<br>**(Yes vs No)** | 0.70 (0.62–0.80) | 0.79 (0.68–0.92) | 0.89 (0.75–1.06) |
| <sup>1</sup>Model 1: adjusted for gender, race, age, education, and ratio of family income to poverty.<br><sup>2</sup>Model 2: adjusted for Model 1 plus total physical activity, smoking, alcohol, and BMI. |  |  |  |

### 3.3. Association of frequency of organic food purchase with hypertension

Compared to never purchasing organic food, the odds of hypertension were (aOR = 0.83, 95% CI: 0.55–1.27) for always or mostly purchasing organic food, (aOR = 0.91, 95% CI: 0.71–1.16) for sometimes purchasing organic food, (aOR = 0.87, 95% CI: 0.67–1.14) for rarely purchasing organic food (**Table 3**).

**Table 3.** Association between frequency of organic food purchase and hypertension, NHANES 2007–2008.

| **Organic food purchase frequency** | **Odds Ratio (OR) 95% CI Crude** | **Odds Ratio (OR) 95% CI Model 1<sup>1</sup>** | **Odds Ratio (OR) 95% CI Model 2<sup>2</sup>** |
| --- | --- | --- | --- |
| **Never** | 1.00 (Reference) | — | — |
| **Rarely** | 0.79 (0.57–1.11) | 0.80 (0.61–1.06) | 0.88 (0.67–1.14) |
| **Sometimes** | 0.71 (0.58–0.87) | 0.78 (0.61–1.00) | 0.91 (0.72–1.16) |
| **Always / Most** | 0.56 (0.39–0.81) | 0.69 (0.46–1.05) | 0.83 (0.55–1.27) |
| <sup>1</sup>Model 1: adjusted for gender, race, age, education, and ratio of family income to poverty. |  |  |  |
| <sup>2</sup>Model 2: adjusted for Model 1 plus total physical activity, smoking, alcohol, and BMI. |  |  |  |

### 3.4. Sensitivity analysis

Sensitivity analysis of organic food purchase and hypertension yielded (aOR = 0.97, 95% CI: 0.82–1.13) (**Supplementary Table 1)**. Compared to never purchasing organic food, the odds of hypertension were (aOR = 0.77, 95% CI: 0.53–1.11) for always or mostly purchasing organic food, (aOR = 0.97, 95% CI: 0.79–1.18) for sometimes purchasing organic food, (aOR = 1.15, 95% CI: 0.84–1.58) for rarely purchasing organic food (**Supplementary Table 2**).

## 4. Discussion

The purpose of this study was to investigate the impact of organic food purchase and its frequency on hypertension in a nationally representative U.S. population. Overall, our findings showed lower estimated odds of hypertension across most models of organic food purchase and purchase frequency, although most estimates were imprecise. Most estimates were imprecise, likely reflecting possible misclassification of organic food purchase frequency, restriction of the frequency of organic food purchase analysis to the 2007-2008 cycle, due to the question not being asked in 2009-2010, not being asked in the questionnaire, and residual confounding. In the sensitivity analysis odds of hypertension were attenuated to the main analysis for both organic food purchase and purchase frequency, though confidence intervals were of similar width.

Organic food is often perceived as a healthier alternative to conventional foods ^28–30^. The evidence for the health benefits of organic food is limited, but existing studies have investigated potential mechanisms of action on blood pressure ^26,31^. These mechanisms can be attributed to strict organic agricultural practices, which limit or prohibit certain practices, including synthetic pesticides, genetically modified organisms, irradiation, antibiotics, and hormones. Several studies have suggested a link between pesticides and blood pressure, with a positive association observed for increased blood pressure for certain pesticides ^15,25,27^. Another mechanistic action could potentially be explained by organic agricultural practices. Organic food may contain higher levels of certain nutrients in produce and animal products ^11–14^. These nutrients have been associated with a reduction in blood pressure ^12,22,32,33^.

Some studies indirectly explored the association between organic food and hypertension. For instance, the KOALA birth cohort study examined the association between organic food consumption during pregnancy and associated health-related characteristics ^16^. Participants who consumed organic food had a lower incidence of hypertension than non-organic consumers; the association with blood pressure didn’t appear linear. Similarly, the NutriNet-Santé study examined the association between organic food consumption and metabolic syndrome (MetS) ^17^. Having a blood pressure ≥130/85 mmHg or taking antihypertensive medication was defined as one criterion for MetS. Results showed that higher consumption of organic food was associated with a lower probability of having MetS.

Nevertheless, associations between organic food purchase and hypertension may reflect health-conscious behavior rather than the direct effect of organic food purchase. Individuals who purchase organic food are more likely to adopt better lifestyle choices and healthier diets ^34,35^. Also, individuals who have higher socioeconomic status, including higher family income and education, were more likely to purchase organic food ^36^. As a result, lower odds of hypertension may be due to confounding of these behaviors and status rather than organic food purchase.

Our study has several strengths in comparison to prior studies. In contrast to the previous two studies ^16,17^, our current study directly estimated the association between organic food and hypertension by clearly defining the outcome of hypertension. Previous studies did not observe the direct effects of organic food on hypertension or focused on specific populations, limiting the generalizability of their results. This study is a nationally representative population and, therefore, more likely to be generalizable to the US population. Using NHANES data, we controlled for potential confounding factors. Finally, our study also investigates the frequency of organic food purchase, allowing us to evaluate dose-response patterns.

Despite these strengths, our study has a few limitations. The study design is cross-sectional, which cannot establish temporality between organic food purchase and hypertension. In fact, there is susceptibility to reverse causality, that is, individuals with diagnosed hypertension may choose to modify their diet and choose to eat organic foods. Given that this is a cross-sectional study, it captures only a mixed diet of organic and non-organic foods, or a fully organic diet. By contrast, a randomized controlled trial could isolate effects on a fully organic diet. Our exposure measurement uses a proxy for organic food consumption and does not reflect actual organic food consumption. There could be individuals whose frequency of organic food purchase doesn’t reflect the actual consumption. Organic food consumption is often associated with healthier dietary habits and lower obesity rates; thus, it is difficult to define the benefits of organic food ^34–36^. Finally, although we have adjusted for potential confounders, residual confounding may still occur.

## 5. Conclusion

Our findings suggest that organic food purchase, a proxy for organic food consumption, may be associated with lower odds of hypertension, though confidence intervals were wide reflecting limited precision in our estimates. These findings may reflect either the true benefits of organic food consumption, including lower pesticide levels and higher nutrient content, or reflect health-seeking behaviors among health-conscious, healthy, and highly educated individuals. Future studies should use longitudinal designs and randomized trials better to understand the effects of organic food on health outcomes.

## Data Availability

The datasets generated and/or analyzed during the current study are available in the [National Health and Nutrition Examination Survey (NHANES)] repository, at [https://wwwn.cdc.gov/nchs/nhanes/default.aspx]

## List of abbreviations

DASH: Dietary Approaches to Stop Hypertension
NHANES: National Health and Nutrition Examination Survey
CVD: Cardiovascular Disease
BP: Blood Pressure
aOR: adjusted odds ratio
HEI: Healthy Eating Index
MEC: Mobile Exam Center
AChE: Acetylcholinesterase
MetS: Metabolic Syndrome

## Decelerations

### Ethics approval and consent to participate

Not Applicable

### Consent for publication

Not Applicable

### Availability of data and materials

The datasets generated and/or analyzed during the current study are available in the [National Health and Nutrition Examination Survey (NHANES)] repository, at [NHANES Questionnaires, Datasets, and Related Documentation]

### Funding

RAN was supported by the National Institute on Minority Health and Health Disparities [K01MD014163], National Institutes of Health, Bethesda, MD. The funder had no role in study design, data collection and analysis, decision to publish, or preparation of the manuscript. The first author (CC) had full access to all the data in the study and takes final responsibility for the paper.

### Author Contributions

**CC** conducted the data analysis and wrote the first draft. **RAN** led the problem definition, conceptualized and supervised the implementation of the analysis, reviewed and revised the manuscript. **OAA** contributed to the interpretation of the results and critically reviewed and revised the manuscript. **LC** contributed to the methodology, reviewed and revised the manuscript. All authors read and approved of the final manuscript.

## Acknowledgements

Not Applicable

## Supplementary Materials

Table S1: Association between organic food purchase and hypertension, NHANES 2007–2008

Table S2: Association between the frequency of organic food purchase and hypertension ((≥ stage 1), NHANES 2007–2008

Figure: Flow chart for participant inclusion and exclusion

