## Supplementary figures and images for "Association Between Purchase of Organic Food and Hypertension Among US Adults: NHANES 2007-2010"

### fig.JPG

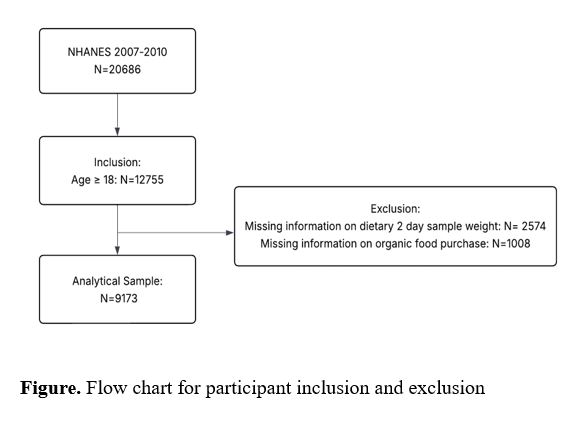

### tbl_s1.JPG

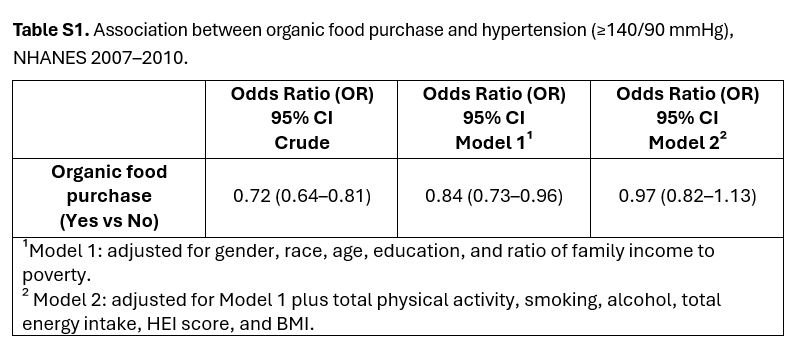

### tbl_s2.JPG

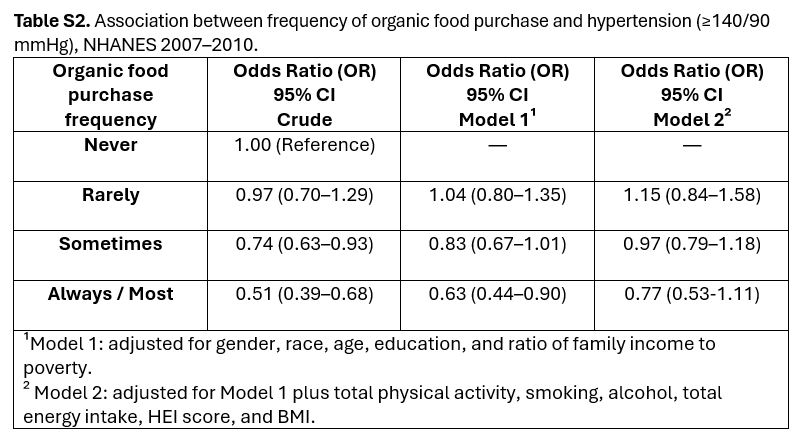
